# Distinct transcriptomic signature of mRNA and microRNA in ulcerative colitis and irritable bowel syndrome

**DOI:** 10.1101/2021.03.19.21253573

**Authors:** Hanqing Luo, Jiacheng Dai, Chunyu Liu, Yue Lee, Hong Lv, Jiaming Qian

## Abstract

**Background:** Ulcerative colitis (UC) and irritable bowel syndrome (IBS) are both chronic bowel diseases involving stress. To identify genes differentially expressed in UC and IBS, and to determine whether psychological stress can influence those gene expressions, we conducted this pilot study.

**Methods:** Patients of UC, irritable bowel syndrome (IBS) and normal controls (NC) (n=5 in each group) were recruited. Perceived stress scale (PSS) was adopted to assess psychological stress level. Sigmoid mucosa samples were collected during colonoscopy. Differentially expressed genes (DEGs) of both mRNA and microRNA (DEG-mRNA and DEG-miRNA) were identified by RNA-Seq and microarray, respectively. Weighted gene co-expression network analysis (WGCNA), gene ontology, and microRNA target analysis were performed to identify regulatory relationships and pathways involved. Pearson correlation was performed to identify the relationship between DEGs and PSS score.

**Results:** In total, 1,770 and 938 DEG-mRNAs, 107 and 3 DEG-miRNAs were identified in UC and IBS (nominal P<0.05), respectively. The transcriptome changes in UC and IBS were highly correlated but transcriptome severity was larger in UC. There were 268 overlapped DEG-mRNAs and 3 overlapped DEG-miRNAs between UC and IBS. Median PSS score was 27.5 (24.7, 40.0) in UC, 27.0 (17.5, 32.5) in IBS and 22.0 (12.0, 28.5) in NC. Five of the UC’s DEG-mRNAs were significantly correlated to PSS scores, including SLC4A7, PLCB1, SPATA33, DECR2, RP11-792A8.4. There were no DEGs correlate with PSS scores in IBS. One module enriched for immunological pathways was upregulated in UC. Metabolic pathways were enriched in IBS. Within the UC-related co-expression module, we identified 18 DEG-mRNAs that were also targets of DEG-miRNA.

**Conclusions:** Integration of microRNA and mRNA expression profiles identified changes of gene expression in both UC and IBS. Although the two diseases have significant overlapped transcriptomic changes, they have distinct signature. Immunological pathways are involved prominently in UC while metabolic pathways in IBS. Psychological stress is uniquely associated with the transcriptome changes of UC.

## BACKGROUND

Ulcerative colitis (UC) is a kind of inflammatory bowel disease (IBD). It is characterized by chronic mucosal inflammation, with symptoms such as diarrhea and rectal bleeding. Irritable bowel syndrome (IBS) is a kind of functional gastrointestinal disease with symptoms including abdominal pain, diarrhea or constipation. Some published literatures have shown that IBD and IBS have overlapped features. Stress is gradually recognized as a factor in the development of UC (1, 2). In IBS, psychological stress plays a major role in the etiology through brain-gut axis (3). However, the etiology of both diseases is unknown.

As genome-wide-associations studies (GWAS) became available, more than two hundred genetic risk loci had been identified for IBD (4, 5). Several genomic regions had also been identified for IBS (6). Transcriptome studies, including both mRNA and microRNA, have contributed to our understanding of IBD and IBS pathogenesis(7, 8). A lot of mRNAs, such as HLA-DRB1 and TAP2, have been found to be differentially expressed in the colon of UC(9). A number of miRNAs, as modulators of gene expression, have been found to be involved in the initiation, development and progression of UC, such as miR-21(10), miR-31(11). In IBS, there were also some mRNA and microRNA found to be differentially expressed, such as INADL (12)and miR-16 (13). However, it is hard to pinpoint exact risk genes for those polygenic diseases. Network analysis may shed new light to the disease mechanisms.

There are studies combining mRNA and miRNA profiles to identified molecular mechanisms in UC (14, 15). The analysis which paired miRNA-mRNA was also used to identify potential candidates in IBS (16). However, few studies compared transcriptomic signature in UC and IBS to date. Therefore, we hypothesize that miRNA-mRNA co-expression network and comparing transcriptome study will lead to better understanding of the core mechanisms of these two diseases with similar symptoms, UC and IBS.

In this study, we aim to compare the transcriptome changes in UC and IBS, and their connections with psychological stress and immune changes to reveal their distinct and shared mechanisms. We performed this pilot transcriptome study with both mRNA and miRNA in UC and IBS. Differential expression analysis and weighted gene co-expression network analysis (WGCNA) were performed.

## METHODS

### 1.1 Patients and Controls

The study protocols were submitted to, and approved by the ethics committee of Peking Union Medical College Hospital (PUMCH). The approval number was S-723. Patients and Controls were prospectively recruited. Ulcerative patients were diagnosed using established criteria (17). IBS patients were diagnosed by ROME III criteria (18). A normal control (NC) was defined as being asymptomatic, undergoing colonoscopy examination for routine check-up and no abnormality was found under colonoscopy. Further criteria were as follows.

Inclusion criteria: age between 16 and 80 years old; UC or IBS diagnosis confirmed by gastroenterology physicians of PUMCH; signed informed consent. Exclusion criteria: past total colorectal resection; comorbidity with severe physical or mental illness, such as heart failure, schizophrenia and major depressive disorder; any other contraindications for colonoscopy; UC patients without the presence of inflammation in the sigmoid colon; self-reported difficulties in reading or understanding.

In this pilot study, 5 UC patients, 5 IBS patients and 5 NC were recruited.

### 1.2 Data and questionnaire collection

Data on demographics (age, gender) and degree of inflammation in UC patients were collected. In UC patients, inflammation of sigmoid mucosa was evaluated according to Mayo scores (19) under colonoscopy. Briefly, active inflammation was scored 0 as no inflammation, 1 as obscure texture of the blood vessel, 2 as loss of texture of the blood vessel, and 3 as ulceration and spontaneous bleeding.

All of the participators were asked to fill a questionnaire to assess their perceived stress in the last month. Version PSS-14 of the Perceived Stress Scale (PSS) (20) was used. PSS-14 is a 14-item questionnaire, focusing on the feelings of stress and handling of stress during the preceding month. The higher the PSS score, the greater the perceived stress. Its psychometric properties have been validated in a Chinese population (21).

### 1.3 Tissue Collection and RNA extraction

Biopsies were taken from the most affected part in sigmoid or from the middle of the sigmoid if there was no evidence of inflammation. A single endoscopist was arranged to perform colonoscopy, take biopsies and assess the degree of inflammation. Biopsies were immediately stored in the RNAlater® (Ambion, Inc., Austin, TX) at 4°C for the first 24 hours, and then frozen and kept at −80°C until RNA extraction.

Total RNA was extracted from frozen biopsies using Trizol® (Invitrogen, Carlsbad, CA) according to a standard protocol. Sample quality was assessed by both a photo-spectrometer (Nano Drop Technologies®, Thermo Scientific, Wilmington, DE) and Agilent 2100 Bioanalyzer. The 260/280 ratios in all samples were between 1.8 to 2.0, and the RNA integrity number (RIN) were ≥7. Total RNA was required to be more than 1μg before miRNA chip examination.

### 1.4 Message RNA Sequencing and small RNA chip examination

Message RNA (mRNA) sequencing library was built for each sample by using KAPA Stranded mRNA-Seq Kit Illumina® platform (KAPA BIOSYSTEMAS). mRNA libraries were sequenced on the HiSeq2500 (Illumina) following the manufacturer’s protocol. The sequencing data generated in this study can be downloaded from the NCBI SRA database at https://www.ncbi.nlm.nih.gov/sra/PRJNA674301.

Affymetrix Genechip miRNA4.0 Array was used to detect the miRNAs. The probe sequences covered all the mature miRNA in the miRbase V20 and small nuclear RNAs (snoRNAs and scaRNAs) of humans. Total RNA was labeled with FlashTag™ Biotin RNA Labeling Kit. After labeling, gene chip hybridization reaction, and scanning, Affymetrix® GeneChip® Command Console® Software (AGCC) was used to save the chip’s fluorescent scanning image into a .DAT file for further analyses. The microarray data can be downloaded from the NCBI GEO database at http://www.ncbi.nlm.nih.gov/geo/ using accession number GSE160900.

### 1.5 Statistical Methods and Bioinformatics

#### 1.5.1 Data preprocessing

For mRNA data, we profiled 57,821 genes from the 15 samples. Expression levels were quantified with counts by Robust Structural Equation Modeling (RSEM). We normalized expression with gene length and used transcripts per million (TPM) as expression value for downstream analyses. Genes were filtered to include those with TPM > 0.1 in at least 20% samples. After filtering, 17,895 genes were remained.

For miRNA data, we profiled 4,575 probes from the 15 samples. The miRNA probes absent in at least 20% samples were filtered and left 852 miRNAs for downstream analyses. mRNA and miRNA data were combined for differential gene expression analysis and gene co-expression network analysis. Log2 transformed data of RNA-seq and miRNA microarray was quantile normalized, separately (22). The expression level of mRNA data and miRNA data is distributed in the same scale. After that, we combined mRNA data and miRNA data in one data matrix for further analysis. Principle component analysis (PCA) plot and clustering plot were evaluated for sample outliers with one sample excluded.

#### 1.5.2 Wilcoxon signed-rank test for differential expression analysis

To detect individual mRNA and miRNAs that differentially expressed in patients and NC, we performed Wilcoxon signed-rank test for differential expression analysis using R. UC samples versus NC and IBS samples versus NC were performed by Wilcoxon signed-rank test. Log2 fold-change (log2FC) of mRNA and miRNA was calculated. Finally, we corrected for multiple testing using false discovery rate (FDR) correction. We compared overlapped differentially expressed genes and miRNAs in two diseases with ggplot2 package. The association between differentially expressed genes (DEGs) and PSS scores were evaluated by Pearson correlation analysis.

#### 1.5.3 Gene co-expression network analysis

To evaluate gene expression relationship of individual genes and miRNAs, we performed Weighted Gene Co-expression Network Analysis (WGCNA) (23). Network analysis was performed with the WGCNA package using unsigned networks. A soft-threshold power of 14 was used for all samples to construct scale-free topology (R^2^>0.8). Networks were constructed using the blockwiseModules function. Modules were summarized by their first principle component (ME, module eigengene), and defined using Pearson correlation. Genes within each module were prioritized based on their module membership (kME), defined as correlation to the module eigengene. Module-disease associations were evaluated using Pearson correlation, with UC and IBS versus controls, respectively. We also correlated modules with age, gender and PSS. The p-value of correlation were adjusted by FDR correction.

#### 1.5.4 Gene ontology analysis

We selected genes from modules which were significant different in disease versus NC, for gene ontology analysis. We used WebGastalt (WEB-based Gene SeT AnaLysis Toolkit) for gene functional enrichment analysis (24).

#### 1.5.5 miRNA target overlap

For miRNA in significant modules, we searched miRNA target gene from miRTarBase (25). We summarized the target genes that also have high kME in the same module.

#### 1.5.6 DEG overlap with GWAS findings

To evaluate whether the DEG has genetic basis, we compared the DEG results and gene co-expression modules with GWAS findings. We downloaded GWAS results of UC and IBS from GWAS Catalog (http://www.ebi.ac.uk/gwas/).

## RESULTS

### Differential expression

One UC patient was excluded because she was an outlier in PCA and cluster analysis (Fig. 1). The PCA plot and clustering result also indicate that there were no outliers among all the samples. The information of participants included in DEG and differentially expressed miRNA analysis was given in table 1.

**Table 1.** The characteristics of participants in final analyses.

|  | UC (n=4) | IBS (n=5) | NC (n=5) |
| --- | --- | --- | --- |
| Male, n | 3 | 3 | 3 |
| Age* | 35.5 (24.3, 47.5) | 38.0 (28.5, 48.0) | 44.0 (40.5, 52.0) |
| PSS score* | 27.5 (24.7, 40.0) | 27.0 (17.5, 32.5) | 22.0 (12.0, 28.5) |

\*Values are median (interquartile range). UC: ulcerative colitis, IBS: irritable bowel syndrome, NC: normal control, PSS: perceived stress scale, n: number.

**Fig. 1.**
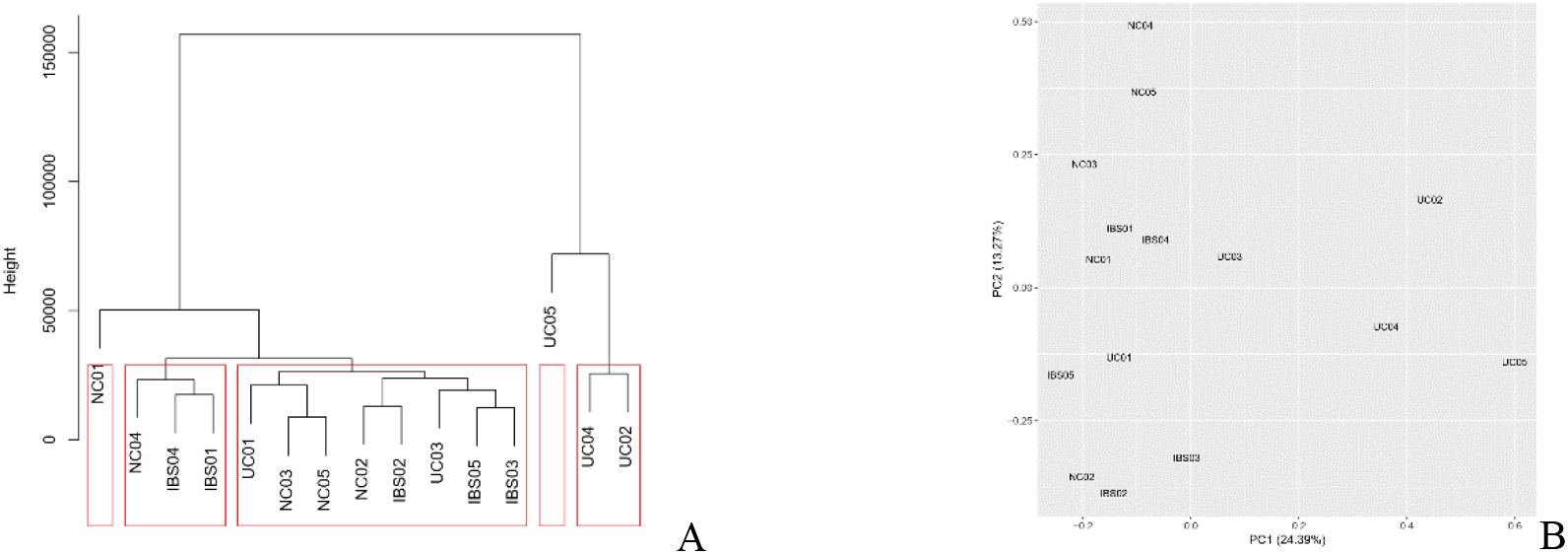
The plot of clustering and principle component analysis (PCA) of all participants. Each participant is represented by each number. A: UC05 was excluded because the height of clustering showed this sample was an outlier. B: UC05 was not gather together with other samples, which was same as clustering result.

Comparing with NC, there were 1770 DEGs and 107 differentially expressed miRNAs in UC patients, 938 DEGs and 3 differentially expressed miRNAs in IBS patients by nominal P<0.05. However, after multiple testing correction, none of the DEGs or differentially expressed miRNAs was significant according to FDR of P<0.05.

Five of the DEGs were related to PSS scores in UC with P <0.01, including SLC4A7 (ρ=0.94), PLCB1 (ρ=0.92), SPATA33 (ρ=−0.90), DECR2 (ρ=−0.88), RP11-792A8.4 (ρ=−0.87). There were no DEGs correlate with PSS scores in IBS.

In UC and IBS, there were 268 overlapped DEGs. Majority of the DEGs (263) had consistent change in the two diseases. Only five genes were upregulated in IBS but downregulated in UC. Among these shared genes, AHR, GPR18, CD28, FAM26F, AC019330.1, LYRM9 are part of GWAS signals in UC. AC019330.1 and LYRM9 were both downregulated in UC and IBS, while others were upregulated in UC and IBS.

The regression slopes of UC and IBS fold change relative to control is 0.85. This plot indicated that these two diseases have many similar gene changes in the same direction, and the transcriptomic severity was larger in UC than in IBS (Fig.2).

**Fig. 2.**
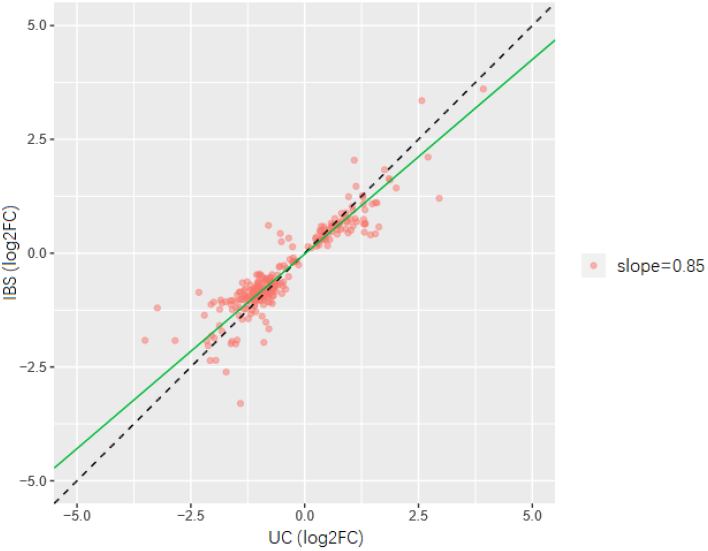
The regression slopes of UC and IBS fold change effect size. The green line shows the slope is 0.85, indicating the transcriptomic severity was larger in UC than in IBS. Values are log2 transformed fold change of differential expressed gene (UC vs control or IBS vs control).

### Gene co-expression network and gene ontology analysis in UC and IBS patients

In the co-expression network analysis, we identified 44 modules (Fig.3). Based on Eigengene differential analysis, Module 6 (FDR-corrected p=0.04) and 11 (FDR-corrected p=0.02) were identified as significantly different in UC versus NC. Module 15 (FDR-corrected p=0.08) and 2 (FDR-corrected p=0.08) were different between IBS and NC (Fig.4). Module 6 and 15 were upregulated in diseases, while module 11 and 2 were suppressed. No significant correlation was detected between any of these modules and PSS scores.

**Fig. 3.**
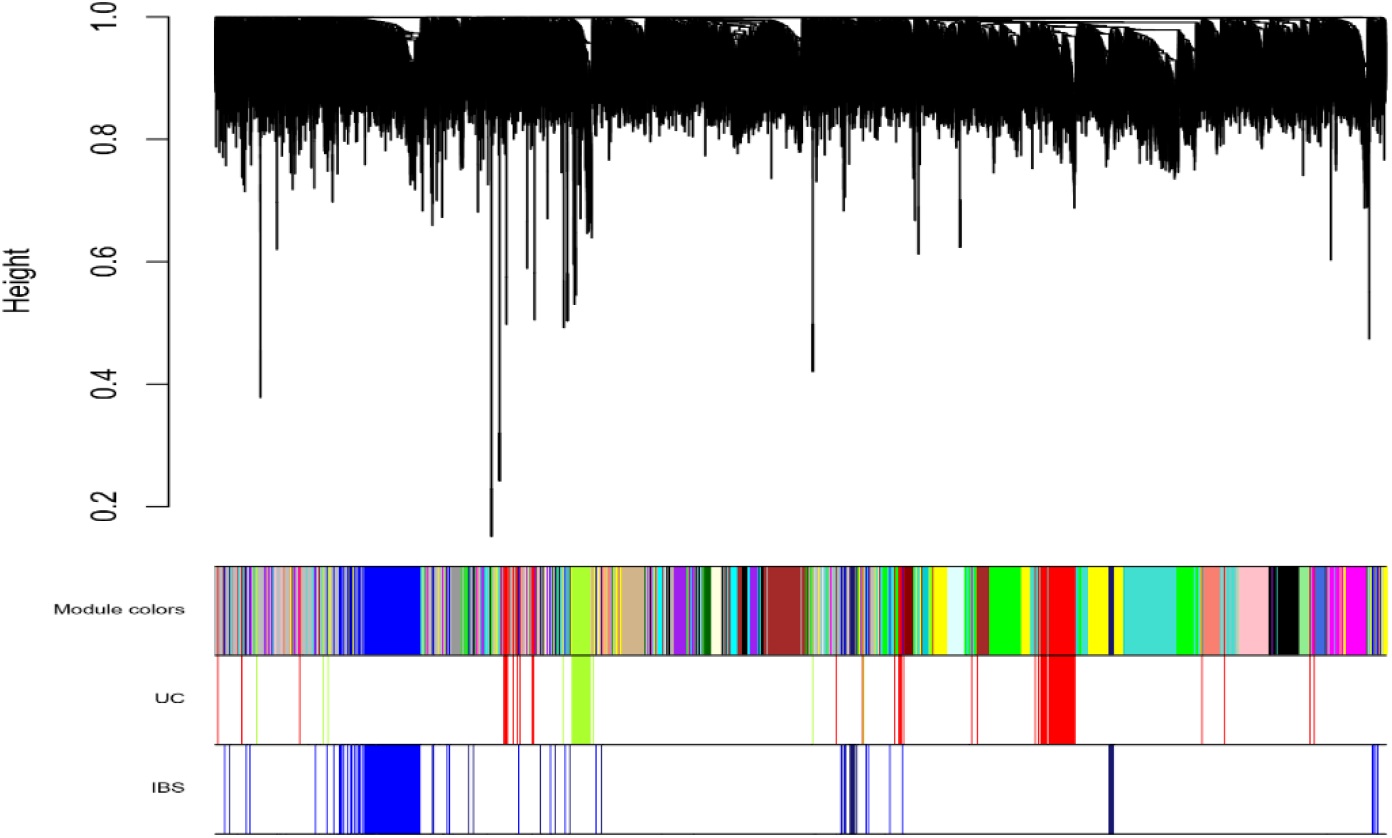
Network dendrogram from co-expression topological overlap of genes in WGCNA. Color bars show gene co-expression modules. Different colors represent different modules. Color bars of UC and IBS represent disease-related gene co-expression module.

**Fig. 4.**
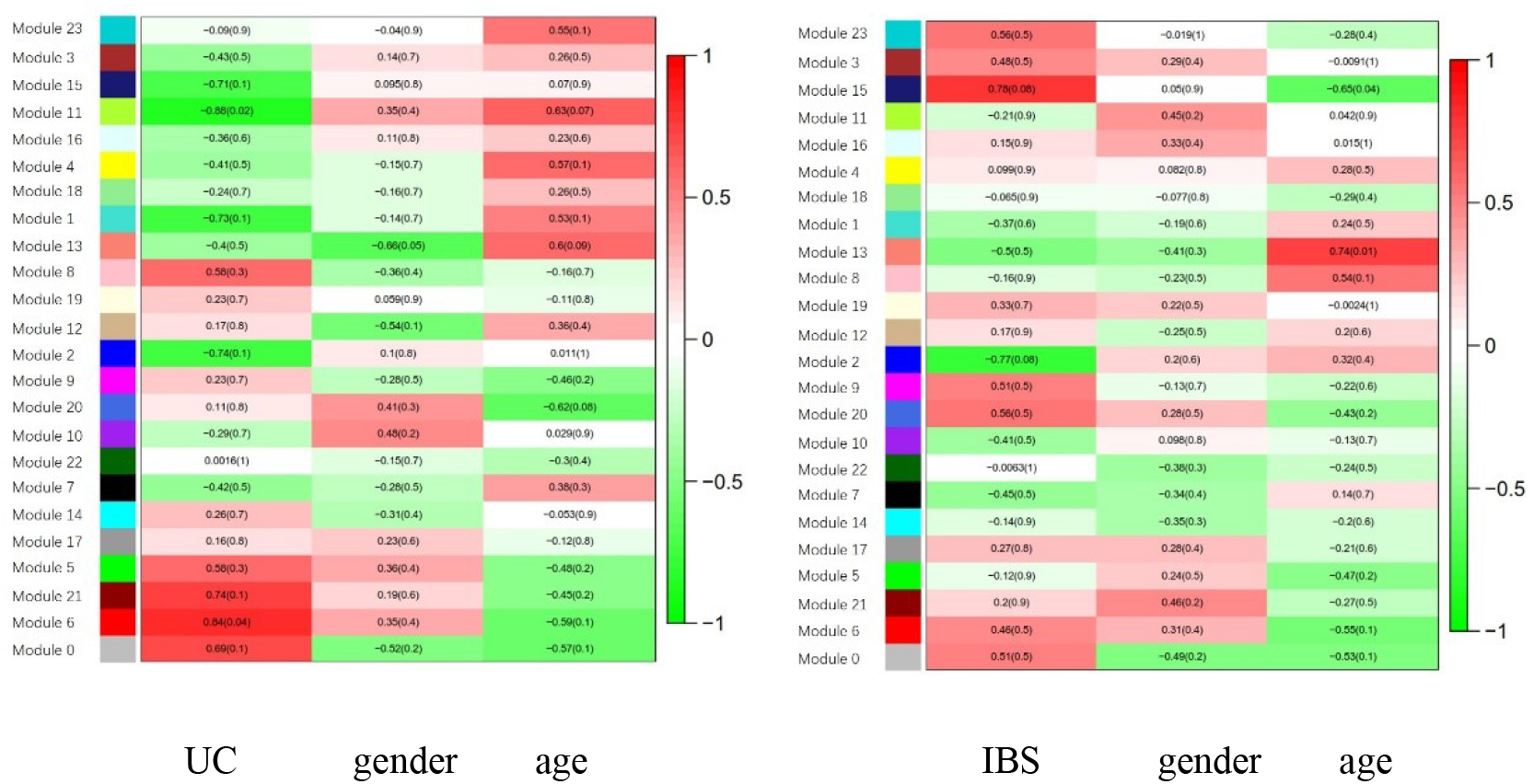
Heatmap of gene co-expression module that correlate to disease state, gender or age of participants. Values are Pearson correlation coefficient and q value (in the bracket) after multiple testing.

In the gene ontology annotation on these disease-associated modules, Module 6 was enriched for activation of immune response, immune response-regulating signaling pathway, leukocyte cell-cell adhesion and regulation of cell activation pathways. Module 15 was enriched for mitochondrial-related pathways. Module 2 was enriched for DNA transcription and DNA metabolic regulation process.

Comparing UC to IBS showed a variety of classes of differential pathway regulation (Table 2). Leukocyte cell-cell adhesion pathway was found elevated in UC, but suppressed in IBS. Immune response is activated in UC. In contrast, pathways annotated as adaptive immune response and inflammatory response to antigenic stimulus were negatively regulated in IBS.

**Table 2.**
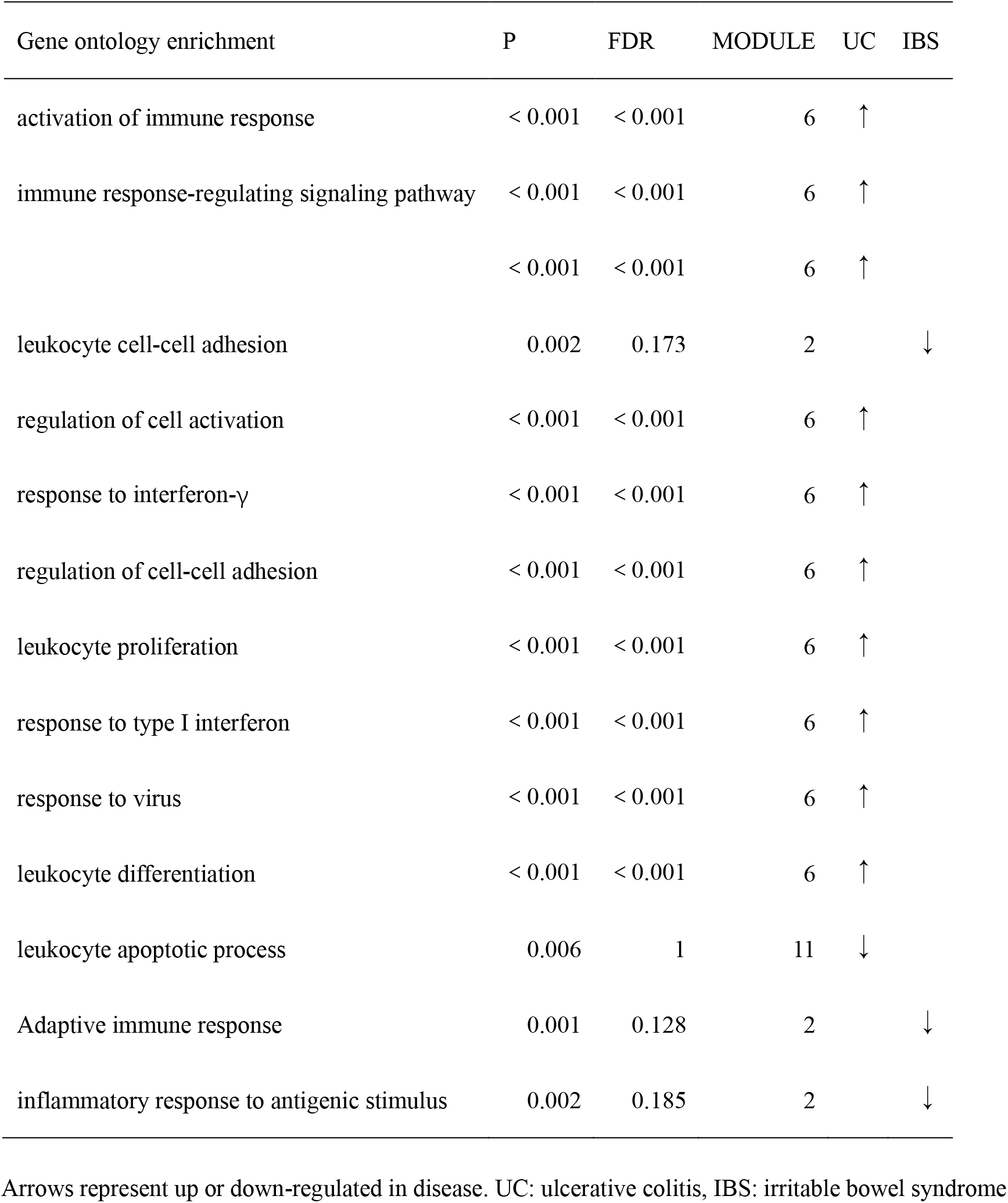
Biological pathways in disease-related modules related to immune responses in UC and IBS.

| Gene ontology enrichment | P | FDR | MODULE | UC | IBS |
| --- | --- | --- | --- | --- | --- |
| activation of immune response | < 0.001 | < 0.001 | 6 | ↑ |  |
| immune response-regulating signaling pathway | < 0.001 | < 0.001 | 6 | ↑ |  |
|  | < 0.001 | < 0.001 | 6 | ↑ |  |
| leukocyte cell-cell adhesion | 0.002 | 0.173 | 2 |  | ↓ |
| regulation of cell activation | < 0.001 | < 0.001 | 6 | ↑ |  |
| response to interferon-γ | < 0.001 | < 0.001 | 6 | ↑ | |
| regulation of cell-cell adhesion | < 0.001 | < 0.001 | 6 | ↑ |  |
| leukocyte proliferation | < 0.001 | < 0.001 | 6 | ↑ |  |
| response to type I interferon | < 0.001 | < 0.001 | 6 | ↑ |  |
| response to virus | < 0.001 | < 0.001 | 6 | ↑ |  |
| leukocyte differentiation | < 0.001 | < 0.001 | 6 | ↑ |  |
| leukocyte apoptotic process | 0.006 | 1 | 11 | ↓ |  |
| Adaptive immune response | 0.001 | 0.128 | 2 |  | ↓ |
| inflammatory response to antigenic stimulus | 0.002 | 0.185 | 2 |  | ↓ |

Arrows represent up or down-regulated in disease. UC: ulcerative colitis, IBS: irritable bowel syndrome

### MiRNA targets overlap with genes in module 6

From WGCNA results, we identified module 6 in UC as a critical module. There were 13 miRNAs in this module. We focused on the miRNA target genes and found 98 target mRNA genes for these 13 miRNAs in module 6. Top 18 of target genes are close to hub of the module 6 (p≤0.0001 & kME>0.85) (Fig. 5).

**Fig. 5.**
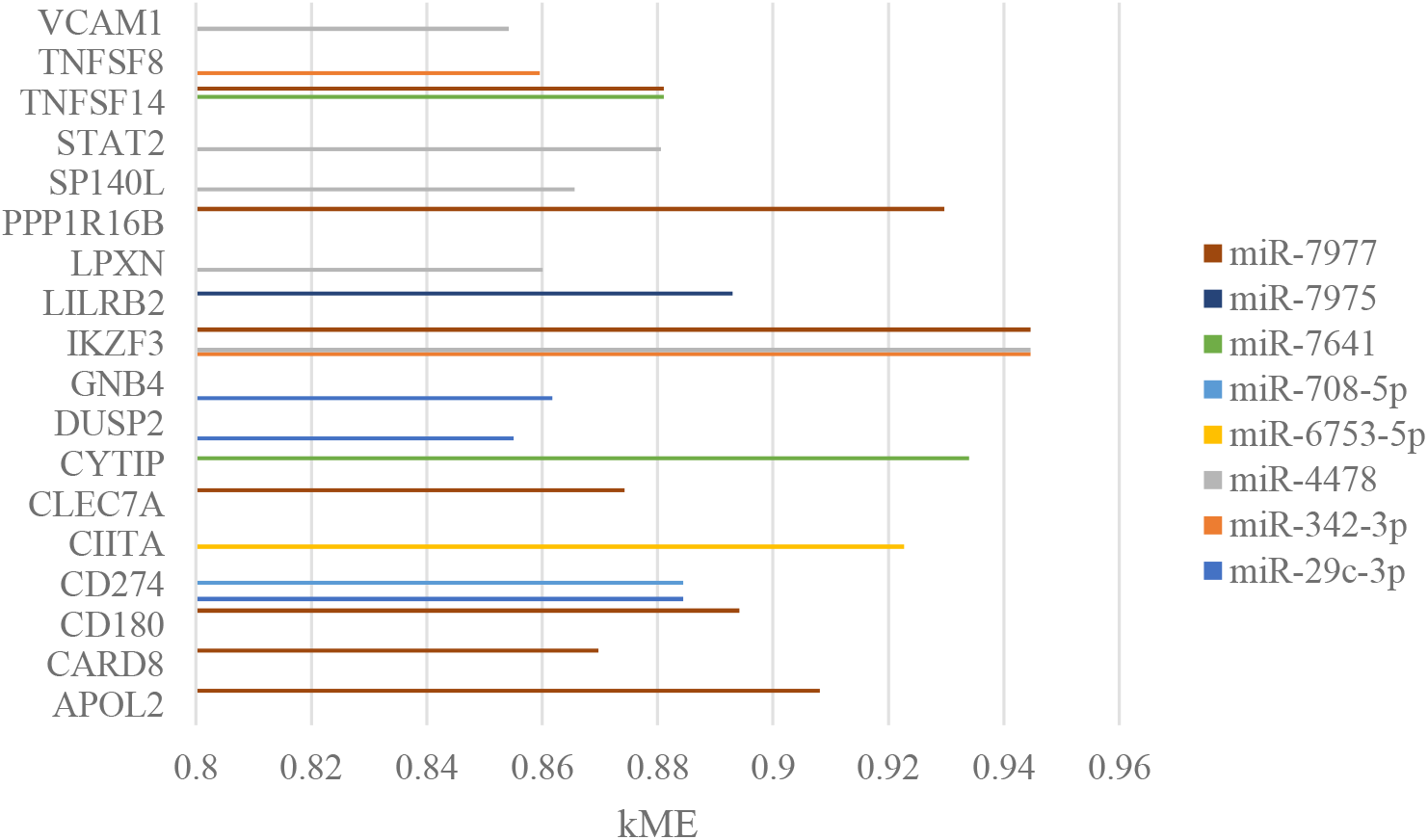
The top gene targeted by high kME miRNA in module 6. Top 18 genes and their kME are provided. Values are module eigengene (kME), which represent the importance of one gene in the module.

### DEG overlap with GWAS findings

Among the 1770 DEGs in UC patients, there were 65 genes overlap with GWAS findings. There were 32 overlapped genes in module 6, such as CIITA, IKZF3, LPXN, TNFSF8. In UC, 5 DEG-mRNAs were part of GWAS signals and shared with IBS. However, they were not overlapped with GWAS signals of IBS. Indeed, we did not find any DEGs overlapped with GWAS findings in IBS.

## DISCUSSION

In this pilot study, even though the sample size is small, the combined miRNA and mRNA expression profiling led to significant findings of upregulated module of gene co-expression and 18 key genes in UC that can survive FDR correction. We found the overall transcriptome changes in UC and IBS were similar with the same gene changes in the same direction, but the transcriptomic severity was larger in UC than in IBS. Psychological stress was only correlated with 5 DEG-mRNAs in UC, but not with co-expression modules or IBS.

Firstly, based on WGCNA and gene ontology analysis, activation of the immune response, immune response-regulating signaling pathway, leukocyte cell-cell adhesion and regulation of cell activation pathways were detected as critical pathways upregulated in UC. The link between immunological process and UC is consistent with previous studies (26). By integrating miRNA and mRNA results, 18 genes were supposed be important in the pathogenesis of UC. In those genes, IKZF3 (27), CIITA (28), TNFSF8 (29), LPXN (30) were also enriched for GWAS signals. IKZF3 is important in the regulation of B lymphocyte proliferation and differentiation (31). Leupaxin (LPXN) can play an inhibitory role in BCR signaling and B cell function (32). That indicates that B cells and adaptive immune system may play a critical role in UC pathogenesis. A former study also identified potential pathways related with immune response in UC with combined data analysis of expression levels of mRNA and miRNA (14). However, the public microarray data of mRNA and miRNA they used were not received from same subjects. Another study used correlation analysis of miRNA and mRNA in 17 UC patients (15) resulted in 319 DEG-mRNAs overlapped with our results, including IKZF3 and TNFSF8. However, compared with correlation analysis, WGCNA used in our study is more efficient (23).

Secondly, similar changes of DEGs were found in UC and IBS, but UC has more DEG and bigger changes. For a long period of time, IBS has been considered as a functional bowel disease without organic lesions. The rise of molecular biology leads to some molecular abnormalities being detected in IBS. Some published literatures have shown that IBD and IBS have overlapped features, such as dysbiosis in gut microbiota and increased mucosal serotonin availability (33). Our results provided important evidence for that shared DEGs may be the basis for the common features of these two diseases. However, UC has bigger changes as an organic disease.

We detected opposite regulatory directions between UC and IBS in some immunological pathways. Immunological pathways were upregulated in UC, which was accordance with other studies (34). However, intestinal immune system seemed to be suppressed in IBS. This finding conflicts with other studies (35). Now that chronic stress can cause immunosuppression (36), immune system might be suppressed in some disease phase of IBS patients. In another way, chemotactic chemokines and pro-inflammatory cytokines were found higher in sera and stool samples of IBS (37). However, the changes in transcriptomic level and protein level may be opposite. Therefore, in IBS, whether immune response is suppressed or activated and whether transcriptome and protein changes in the same direction need further research.

Thirdly, our results also give a glimpse on a possible link between psychological factors and UC. PSS scores were higher in UC patients compared with controls. We found 5 DEG-mRNAs correlated with PSS scores in UC. Among the 5 DEG-mRNAs, SLC4A7 was also upregulated in IBS. As a member of solute carrier family, SLC4A7 encodes a sodium bicarbonate cotransporter serving as an ion channel in the epithelium. The activity of SLC4A7 is modulated by cystic fibrosis transmembrane conductance regulator (CFTR). Mutations in CFTR can lead to channelopathies with diarrhea (38). Ion channels may be important in gastrointestinal function and could be influenced by perceived stress.

PSS scores were also higher in IBS patients. However, we did not detect any DEGs related with perceived stress in IBS. On the one hand, different subtypes of IBS, including constipation-predominant IBS (IBS-C) and diarrhea-predominant IBS (IBS-D), have different degree of correlation with psychological factors (39). In Martínez C’s study (16), they found significant relationship between miRNA-mRNAs expression and PSS scores in patients with IBS-D. A mixed patient group in this study may minimize the significance. On the other hand, psychological stress may regulate function of those genes, rather than directly impact on the expression level.

IBS had co-expression modules enriched for metabolic pathways, such as mitochondrial transport, mitochondrial membrane organization and mitochondrial translation. Mitochondrial function is a key player in coordinating cellular metabolism, immunity and stress response (40). Our results supported that mitochondrial related pathways might function as a core in the complex mechanisms of IBS.

In summary, we performed a pilot study on gene expression and miRNA profiles of UC and IBS, and simultaneously utilizing WGCNA and miRNA target gene overlapping methods to identify core genes. The immunological pathways in UC were highlighted again. Mitochondrial related pathways were emphasized in IBS. Some immunological pathways seemed to play an inverse role between UC and IBS.

Our research has some limitations. Primarily, as a pilot study, the sample size is small, leading to the insufficient statistical power to identify significant DEGs when stringent criteria applied. Nevertheless, we identified reasonable key genes by conducting WGCNA and miRNA analysis. Second, PSS may not precisely reflect the impact of stress on UC patients. In future study, objective testing for physiological markers of stress, such as salivary cortisol levels could be used. Third, due to the limitation of experimental cost, those key genes and pathways were not verified by a second technique and functional experiments. We also do not have another data to validate the overall findings. Additional experiments are warranted to confirm the findings of this study.

## CONCLUSIONS

Overall transcriptome changes in UC and IBS were similar, but the transcriptomic severity was larger in UC. By WGCNA and gene ontology analysis, it was indicated that immunological mechanism might play a central role in UC’s development, while mitochondrial related pathways might be important in IBS. There was some sort of association between perceived stress and UC.

## Data Availability

The sequencing data generated in this study has been deposited in the NCBI SRA database (https://www.ncbi.nlm.nih.gov/sra/PRJNA674301).

The microarray data can be downloaded from the NCBI GEO database at http://www.ncbi.nlm.nih.gov/geo/ using accession number GSE160900.

## LIST OF ABBREVIATIONS

UC: Ulcerative colitis
IBS: Irritable bowel syndrome
IBS-C: Constipation-predominant IBS
IBS-D: Diarrhea-predominant IBS
PSS: Perceived stress scale
DEG: Differentially expressed gene
mRNA: Message RNA
miRNA: microRNA
WGCNA: Weighted gene co-expression network analysis
GO: Gene ontology
GWAS: Genome-wide-associations studies
IBD: Inflammatory bowel disease
HLA-DRB1: Human leukocyte antigen-DR beta 1
TAP2: Transporter 2, ATP binding cassette subfamily B member
INADL: InaD-like gene
PUMCH: Peking Union Medical College Hospital
NC: Normal control
RSEM: Robust Structural Equation Modeling
TPM: Transcripts per million
PCA: Principle component analysis
Log2FC: Log2 fold-change
FDR: False discovery rate
ME: Module eigengene
WebGastalt: WEB-based Gene SeT AnaLysis Toolkit
IQR: Interquartile range
SLC4A7: Solute carrier family 4 member 7
PLCB1: Phospholipase C Beta 1
SPATA33: Spermatogenesis Associated 33
DECR2: 2,4-Dienoyl-CoA Reductase 2
RP11-792A8.4 AHR: Aryl Hydrocarbon Receptor
GPR18: G Protein-Coupled Receptor 18
CD28: CD28 Molecule
FAM26F: Family With Sequence Similarity 26, Member F
AC019330.1 LYRM9: LYR Motif Containing 9
VCAM1: Vascular Cell Adhesion Molecule 1
TNFSF8: Tumor Necrosis Factor (Ligand) Superfamily, Member 8
TNFSF14: Tumor Necrosis Factor (Ligand) Superfamily, Member 14
STAT2: Signal Transducer And Activator Of Transcription 2
SP140L: SP140 Nuclear Body Protein Like
PPP1R16B: Protein Phosphatase 1 Regulatory Subunit 16B
LPXN: Leupaxin
LILRB2: Leukocyte Immunoglobulin Like Receptor B2
IKZF3: IKAROS Family Zinc Finger 3
GNB4: G Protein Subunit Beta 4
DUSP2: Dual Specificity Phosphatase 2
CYTIP: Cytohesin 1 Interacting Protein
CLEC7A: C-Type Lectin Domain Containing 7A
CIITA: Class II Major Histocompatibility Complex Transactivator
CD274: CD274 Molecule(同时也是 PD-L1)
CD180: CD180 Molecule
CARD8: Caspase Recruitment Domain Family Member 8
APOL2: Apolipoprotein L2
CFTR: Cystic fibrosis transmembrane conductance regulator

## DECLARATIONS

### Ethics approval and consent to participate

The study protocols were submitted to, and approved by the ethics committee of Peking Union Medical College Hospital (PUMCH). The approval number was S-723. Informed consent was obtained from all subjects or, if subjects are under 18, from a parent. All methods were carried out in accordance with relevant guidelines and regulations along with the approval.

### Consent for publication

Not applicable

### Authors’ contributions

Han-Qing Luo performed the experiments and drafted the manuscript; Jia-Cheng Dai helped to analyze the data and prepare the figures; Chun-Yu Liu and Jia-Ming Qian edited and revised the manuscript; Han-Qing Luo, Yue Lee, Hong Lv and Jia-Ming Qian designed the research. All authors read and approved the final manuscript.

## Acknowledgements

Not applicable

